# Preferential observation of large infectious disease outbreaks leads to consistent overestimation of intervention efficacy

**DOI:** 10.1101/2020.11.02.20224832

**Authors:** Jon Zelner, Nina Masters, Kelly Broen, Eric Lofgren

**Affiliations:** Dept. of Epidemiology, University of Michigan School of Public Health; Center for Social Epidemiology and Population Health (CSEPH), University of Michigan School of Public Health; Paul G. Allen School for Global Animal Health, Washington State University; Dept. of Math and Statistics, Washington State University

## Abstract

Data from infectious disease outbreaks in congregate settings are often used to elicit clues about which types of interventions may be useful in other facilities. This is commonly done using before-and-after comparisons in which the infectiousness of pre-intervention cases is compared to that of post-intervention cases and the difference is attributed to intervention impact. In this manuscript, we show how a tendency to preferentially observe large outbreaks can lead to consistent overconfidence in how effective these interventions actually are. We show, in particular, that these inferences are highly susceptible to bias when the pathogen under consideration exhibits moderate-to-high amounts of heterogeneity in infectiousness. This includes important pathogens such as SARS-CoV-2, influenza, Noroviruses, HIV, Tuberculosis, and many others

## Introduction

The SARS-CoV-2 pandemic has brought the implications of heterogeneous infectiousness and susceptibility for outbreak control to the forefront of public and scientific conversations around control of the COVID-19 pandemic (1,2). During the pandemic, outbreaks in congregate settings including cruise ships, jails, prisons (3), nursing homes, and long term care facilities have become key foci of SARS-CoV-2 infection and mortality. Superspreading, in which a small fraction of cases drives the bulk of transmission is common in such outbreaks. This is often the result of a confluence of factors in congregate settings including high rates of contact between individuals within the facility, concentration of highly susceptible individuals in a single location, and high viral load in a subset of infectious individuals. Data from such contexts are often used to elicit clues about which types of interventions may be useful in other facilities, using before-and-after comparisons in which the infectiousness of pre-intervention cases is compared to that of post-intervention cases and the difference is attributed to intervention impact. For example, this approach was used in an early and widely-cited paper suggesting that public health countermeasures during the SARS-CoV-2 outbreak on the *Diamond Princess* cruise ship was able to drive the on-board *R*_0_ down from a value of nearly 15 to under 2, potentially preventing more than 2,500 cases (4). In this analysis, we explore the sensitivity of such inferences to the presence of heterogeneity in infectiousness, ranging from the moderate, exponential variation associated with classic SIR models to the significant overdispersion characteristic of superspreading.

### Heterogeneity in SARS-CoV-2 transmission

Analyses dating to the beginning of the COVID-19 pandemic suggest that a large proportion of SARS-CoV-2 transmissions are likely attributable to a relatively small number of infectious individuals, with one study from Hong Kong suggesting that 20% of cases were responsible for 80% of observed transmissions(5), and similar dynamics observed in transmission in the U.S. state of Georgia (6). In a seminal analysis, Lloyd-Smith et al. (7)demonstrated how such heterogeneity in infectiousness was responsible for less-frequent but more explosive outbreaks characterized by high-yield super-spreading events (SSEs). This and other similar observations have led to suggestions that SARS-CoV-2 infection control should focus primarily on identifying the most infectious individuals and screening, isolating, and treating their exposed contacts. Despite the clear importance of understanding which interventions are effective at preventing and slowing SARS-CoV-2 transmission in congregate settings, little attention has been paid to the practical challenges of evaluating the efficacy of interventions meant to shorten or cut the ‘long tail’ of overdispersed transmissibility characteristic of the transmission of SARS-CoV-2 and other acute infectious diseases in congregate settings.

In addition to well-documented heterogeneity in SARS-CoV-2 transmission, extensive variation has also been documented in the transmission of a wide variety of pathogens, including SARS-CoV-1 (8), Ebola (9), noroviruses (10), tuberculosis (11,12), and measles (13,14). Under the best-case scenario, superspreading is known to pose significant challenges for both the efficacy of intervention and the measurement of key transmission parameters (15,16). It has been noted before that selection bias is likely to compound these challenges, but this question has not been formally addressed.

### Outbreaks cause interventions

Infectiousness heterogeneity poses numerous problems for infectious disease surveillance and control. In this analysis we focus in on the specific problem of observation bias. Simply put, outbreaks resulting in the application of interventions are likely to be significantly larger than those that do not. If we observe and analyze only the set of outbreaks requiring intervention, we are likely to overgeneralize the characteristics of this selected set of large outbreaks to the population as a whole. As the average value of the effective reproduction number, *R*, for a given pathogen at the population level declines, due to changing behavior, or the accumulation of naturally or vaccine-derived immunity in the population, analyzed outbreaks will necessarily become progressively less representative of the population as a whole, particularly for those pathogens exhibiting significant variation in infectiousness.

### Pre-post comparisons may lead to erroneous conclusions

Interventions meant to arrest an ongoing outbreak are necessarily *reactive* in nature: An anomalous transmission pattern is detected, and interventions are applied in an effort to stop the outbreak ‘in its tracks’. Intervention effects are often measured using some kind of pre-post comparison, measuring change in average infectiousness prior to and after the institution of interventions. Previous work has shown that even with limited heterogeneity in infectiousness, such before-and-after comparisons routinely lead to overly optimistic estimates of intervention impact (17). This is because analyses of large outbreaks characterized by overt morbidity and mortality are far more likely than analyses of small, non-fatal outbreaks. Consequently, there is typically no pool of similar, uncontrolled outbreaks that can be used as controls to isolate intervention effects. Figure 1 illustrates how this bias towards analyzing the most severe outbreaks induces a causal linkage between the infectiousness of early cases and the application of a treatment. In other words, because interventions are not randomly assigned to outbreaks, there is a significant risk of *confounding by indication* (18) due to systematic differences between outbreaks receiving intervention and those that do not.

**Figure 1:**
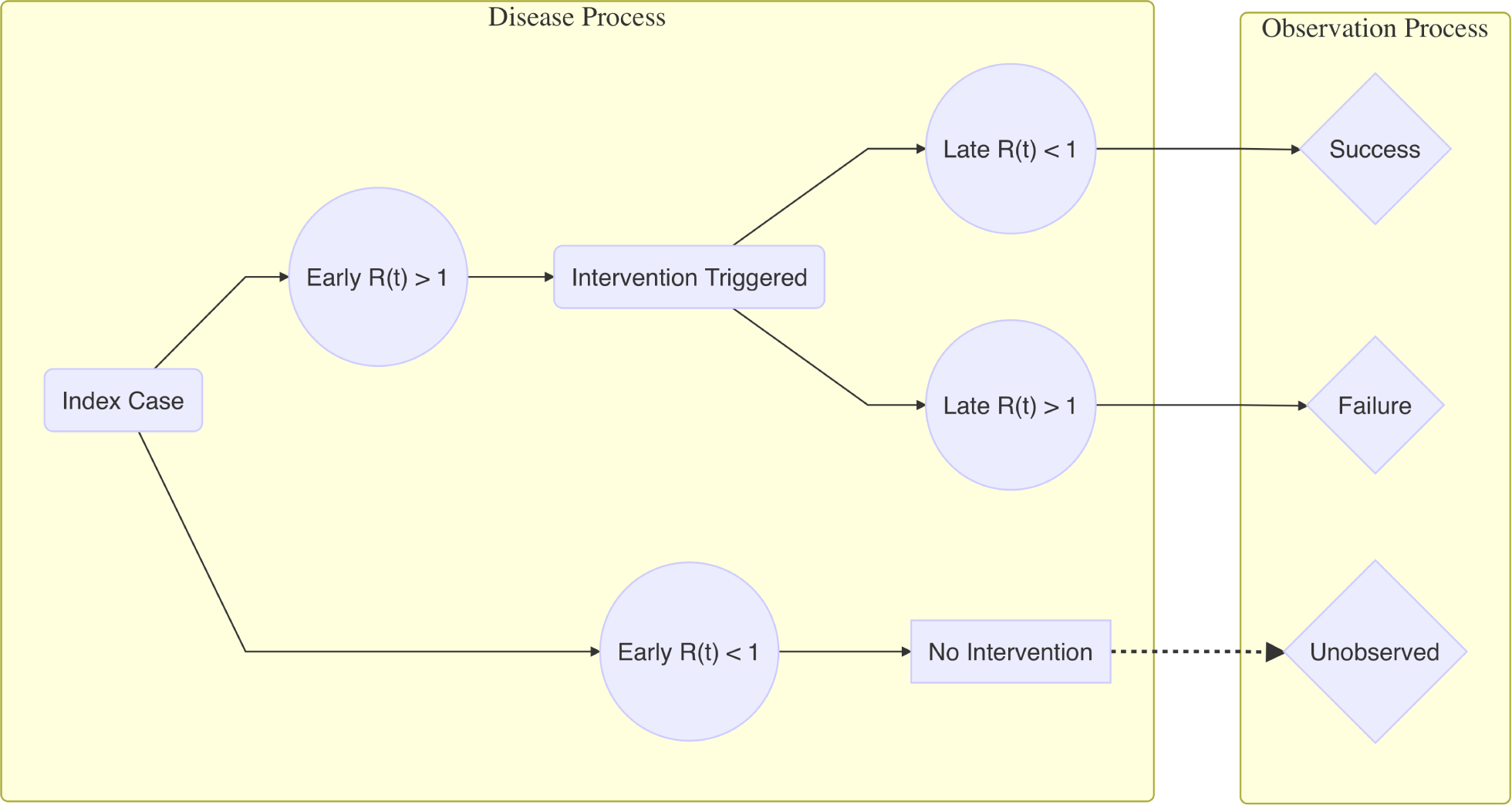
Flow diagram illustrating causation of observation by intervention.

Nevertheless, the pre/post approach continutes to be applied to the measurement of intervention effectiveness for a range of pathogens, including noroviruses (19). SARS (20,21), Hand, Foot, and Mouth Disease (22), pandemic influenza (23), Ebola (9), plague (24), and others. It is important to note that the results of many of these analyses are presented with the caveat that the effects of intervention may be difficult to disentangle from inherent variation in individual infectiousness, contact, etc. Nevertheless, for emerging infections such as SARS-CoV-2, there is often little choice but to rely on such pre/post comparisons. Our goal in the present analysis is to use a transmission modeling approach to identify the situations in which such comparisons are most prone to bias, and to quantify the direction and magnitude of these biases.

## Methods

For clarity, we will make a distinction between the population-level basic reproduction number, *R*_0_, which represents transmissibility throughout the entire population in the absence of immunity or intervention, and the setting-specific transmissibility, denoted *R*_*s*_. This indicates the average transmissibility in a given setting, *s*, such as a jail, school, or nursing home, and may include the impact of measures such as social distancing that may reduce infectiousness, as well as accumulated immunity from natural infection and a potential vaccine.

Before-and-after effect estimates are typically made by estimating 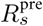 before, and 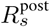 after the intervention. Implicit in this approach is an assumption that: 1) outbreaks selected for intervention and analysis are representative of the population of potential outbreaks, i.e. that 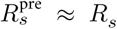, and that 2) 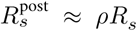, where 0 < *ρ* < 1 is the reduction in transmissibility conferred by the intervention. Put another way, this framework implicitly assumes that the time-evolution of *R*_*s*_(*t*) is primarily a function of intervention, with values of an individual’s infectiousness in setting *s, r*_*i,s*_ representing simple random samples from the setting-specific distribution of infectiousness. In our analysis, we adapt the analytic approach in (17) to understand the implications of overdispersed, heterogeneous infectiousness for the evaluation of reactive interventions following the appearance of an anomalous cluster of cases.

### Transmission model

We use the approach developed by Zelner et al. to model heterogeneous infectiousness in a large norovirus outbreak (10), which builds on the framework of Lloyd-Smith et al. (7). As described above, each case has an *individual* reproduction number represented by a random variable, *r*_*i,s*_ which follows a gamma distribution with dispersion parameter *k*_*s*_ and mean *R*_*s*_, i.e. *r*_*i,s*_ ∼ Gamma(*k*_*s*_, *R*_*s*_/*k*_*s*_) where *R*_*s*_/*k*_*s*_ is equivalent to the shape parameters *θ*_*s*_. This allows us to hold the expected value of *R*_*s*_ across all infectious individuals in a given setting constant while manipulating the variance: As *k* → 0, the variance of individual infectiousness increases towards infinity, and as *k* → ∞, the variance of *r*_*i*_ → 0, i.e. all cases will have exactly the same infeciousness.

To allow for susceptible depletion, we consider a finite-population stochastic SIR model. To account for between-individual variation in infectiousness in the context of a mass-action model, we take advantage of the fact that the distribution of a sum of *N* gamma random variables with shape *k* and scale *θ* is Gamma distributed with shape *kN*. So, we draw the total force of infection on each day, *λ*_*s*_(*t*) = ∑_*i*_ *ℐ*(*τ*_*i*_ = *t*)*r*_*i,s*_ from a Gamma distribution with shape parameter *k*_*s*_*I*_*s*_(*t*) and scale parameter *R*_*s*_/*k*_*s*_, where *τ*_*i*_ is the onset time for individual *i*. So, the risk of infection to each susceptible individual on day *t* is equal 1 − *exp*(−*λ*(*t*)/*N*). We also use this value to estimate the value of *R*_*s*_(*t*) = *λ*(*t*)/ ∑_*i*_ *ℐ*(*τ*_*i*_ = *t*).

Modeled outbreaks are started at *t* = 1 with the introduction of a single infectious individual into a population of 1000 people. Contact is assumed to be density dependent, i.e. the daily per-capita infectiousness of each case 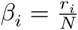. For simplicity, we assume that individuals are infectious only on the day they develop symptoms. Individuals infected on day *t* become infectious on day *t* + 1. We do this because models that estimate *R*_*s*_(*t*) typically report the mean infectiousness of cases with onset at day *t*. We utilize an SIR model rather than an SEIR model because of the potential for pre-symptomatic infectiousness among SARS-CoV-2 cases, and because the inclusion of a latent phase does not impact the overall outbreak dynamics, which are the focus of the current analysis.

### Distinguishing regression toward the mean from intervention effects

We hypothesize that *observed* outbreaks with true setting-specific reproduction numbers, *R*_*s*_ close to one will be disproportionately likely to exhibit larger-than-expected values of *R*_*s*_(*t*) in their early phases, followed by regression toward the mean value of *R*_*s*_ as the outbreak proceeds. When this coincides with the time an outbreak crosses a size threshold, *c*, that makes the outbreak severe enough to trigger intervention, the estimated protective effects attributed to the intervention may be large even when the intervention is completely ineffective.

Concretely, if the value of *R*_*s*_ < 1 in the absence of intervention, on average, the introduction of an infectious individual would be expected *a priori* to not result in an outbreak. When outbreaks of size *z* > *c* occur, the sample mean of daily reproduction numbers prior to crossing the intervention threshhold is will necessarily be greater than the mean at the beginning of the outbreak, outbreak size threshholds *c* > 1. This opens up the risk of what Gelman and Carlin term Type M errors of *magnitude* in analyses in which the estimator of a protective effect of an intervention is the change in the expected value of *R*_*s*_(*t*) before and after intervention. By conditionining on observing only outbreaks with final size *z* ≥ *c*, we increase the likelihood of observing values of Δ*R*_0_ that can be explained by regression toward the mean rather than the causal impact of intervention.

### Simulation strategy

To measure the probability that Δ*R*_0_ will cross unity in the absence of intervention, i.e. the risk of misattribution of outbreak effects to regression toward the mean (RTM), we constructed a design analysis (25) with the following steps:

1. Select parameters *θ* = *c, R*_0_, *k*.
2. Introduce 1 infectious individual with *r*_*i*_ drawn uniformly at random from the true distribution of *r*_*i,s*_into a closed population of 1000 susceptible individuals.
3. Simulate the transmission process over a 14-day period 5000 times for each input parameter set.

Then, for the subset of simulated outbreaks with final size above the outbreak threshold, i.e. *z* > *c*:

1. Determine the day, *ζ* when the intervention threshold is crossed.
2. Calculate 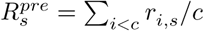 and 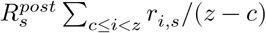.
3. Estimate 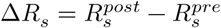.
4. Calculate the proportion of outbreaks in which 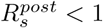 and 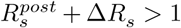. This is the fraction of outbreaks in which a post-intervention drop in *R*_*s*_ which would be attributed to intervention is likely the result of RTM. This value is denoted 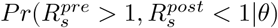.

In all simulations, we explore a range of *R*_*s*_ values from 2.5, representing the rates of uncontrolled SARS-CoV-2 transmission in a completely susceptible population, to values < 1, suggestive of largely controlled transmission characterized by sporadic outbreaks.

## Results

For misattributation of the impact of RTM to intervention effects to occur, the average value of *r*_*i,s*_ prior to the triggering of interventions necessarily must be significantly greater than the mean value of *R*_*s*_ across all outbreaks. Figure 2 illustrates that this is the case across all levels of *R*_*s*_ and *k* included in our simulations, with the ratio of the observed 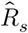 to the true *R*_*s*_ greatest for small values of *k*, with these differences becoming even more pronounced for higher values of *R*_*s*_.

**Figure 2:**
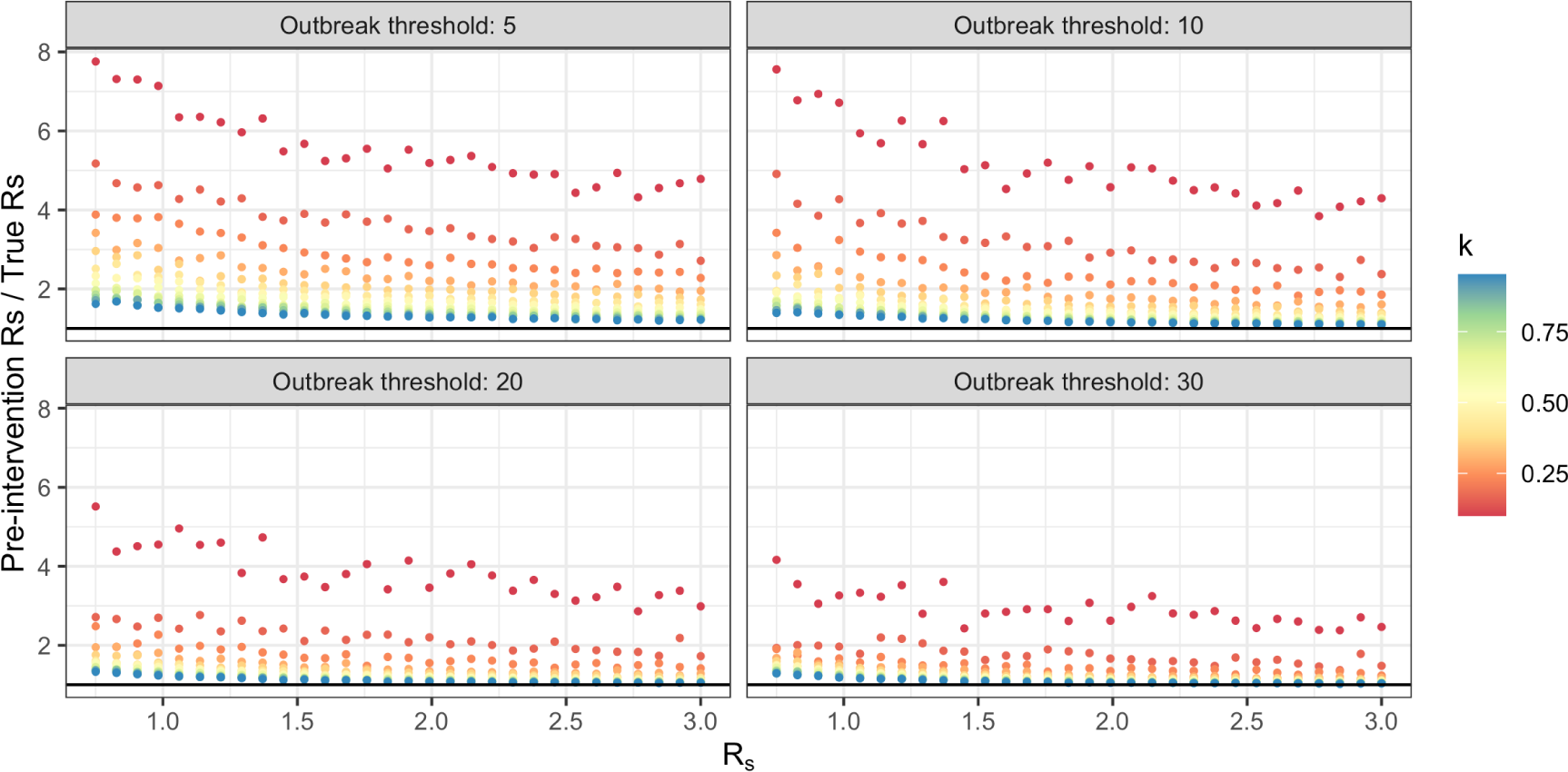
Ratio of pre-intervention to true value of *R*_*s*_. Results are presented as a function of *R*_*s*_ and *k*_*s*_ for outbreak threshholds of 5, 10, 20 and 30 cases

Figure 3 illustrates how these over-estimates may translate into large drops in *R*_*s*_ indicative of effective intervention may occur even if the true value of *R*_*s*_ is above 1, as in the analysis of the *Diamond Princess* SARS-CoV-2 outbreak in (4). Somewhat unintuitively, these effects are most pronounced for higher outbreak thresholds, greater heterogeneity, and larger values of *R*_*s*_. These large drops and higher average pre-intervention infectiousness estimates can be explained by the fact that smaller outbreaks, i.e. those just above the threshold, are more common than very large ones and are more likely to be driven by a small number of early cases with disproportionately high infectiousness.

**Figure 3:**
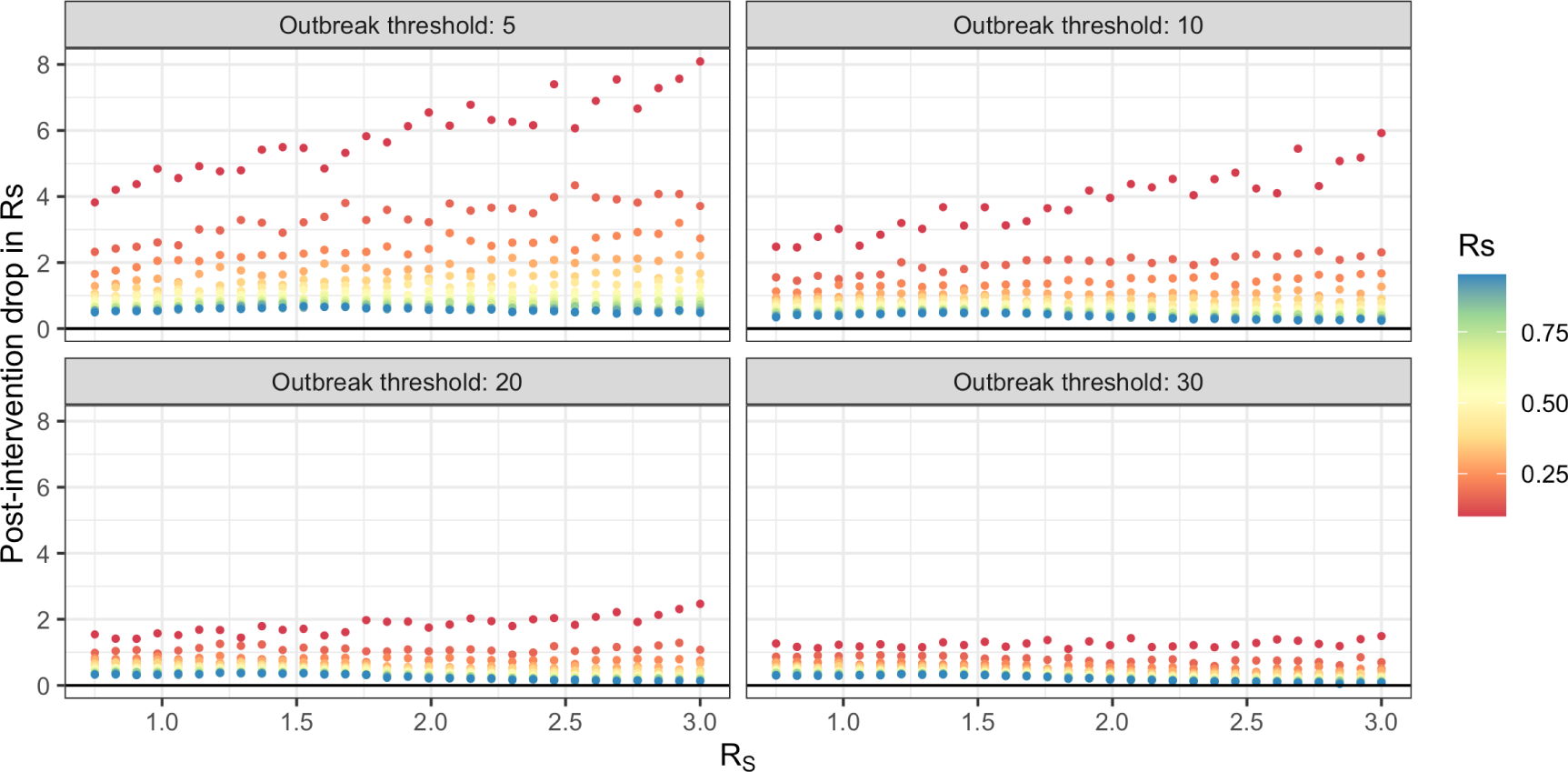
Average decline in R_s_ after crossing a pre-specified intervention threshhold. Results are presented as a function of *R*_*s*_ and *k* for outbreak threshholds of 5, 10, 20 and 30 cases.

Finally, Figure 4 shows the proportion of outbreaks in which 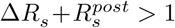, i.e. when a drop in *R*_*s*_ (*t*) suggestive of interventions contributing to the end of the outbreak is likely to be misattributed to regression toward the mean. This shows that for small values of the dispersion parameter - indicative of wide heterogeneity in infectiousness - the expected value of the setting-specific reproduction number can exhibit drops suggestive of effective intervention even when no intervention is applied > 50% of the time, and this is true even when *R*_*s*_ > 1. When the value of *R*_*s*_ is near the the critical value of 1, the probability of misattributation approaches 100% of outbreaks regardless of the outbreak threshhold used. For values of 0.5 < *R*_*s*_ < 1, the probability of misattribution increases with *R*_*s*_. This is a result of the very small probability of drawing a sequence of *r*_*i,s*_ values that sum to greater than 1, with any outbreaks with values of *R*_*s*_ < 1 attributable to stochasticity in the transmission process rather than in the distribution of individual infectiousness. Notably, the outbreak size threshold has little effect on any of these results.

**Figure 4:**
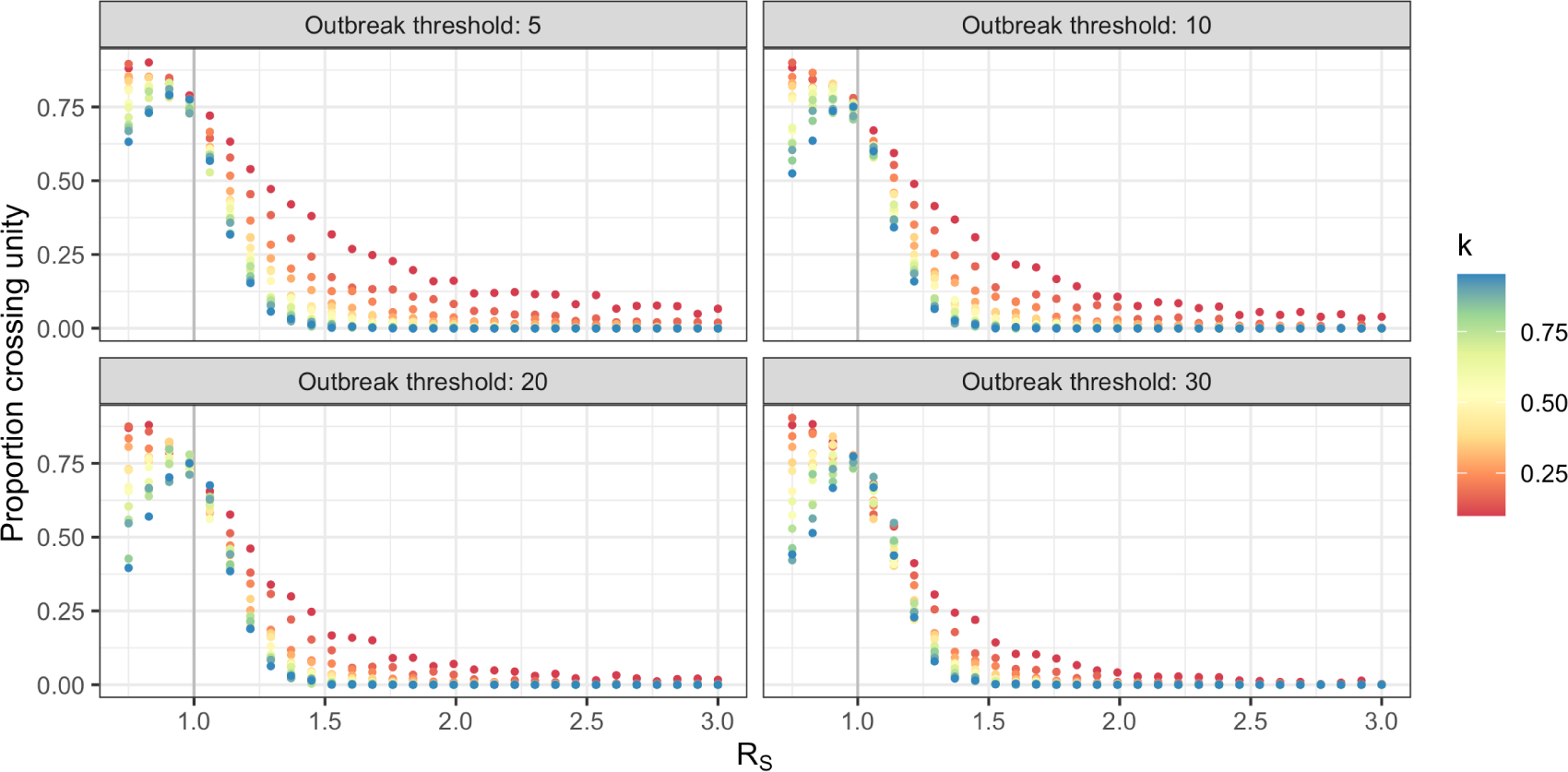
Probability of misattributing the end of an outbreak to interventions. Results are presented as a function of *R*_*s*_ and *k* for outbreak threshholds of 5, 10, 20, and 30 cases

## Discussion

Although our results are schematic in nature, they illustrate key challenges to estimating the impact of interventions on the course of outbreaks of acute pathogens such as SARS-CoV-2 in congregate settings. The finding that increasing amounts of heterogeneity are associated with misattribution of efficacy to ineffective interventions shows that the results of analyses by Cooper et al. (17) are only strengthened in the context of superspreading.

Another implication of these results is that cross-outbreak inferences must be made with significant care: The erroneous conclusion that an intervention was effective in an outbreak in which the true value of *R*_*s*_ was < 1 does not have direct implications for that outbreak, which was in truth self-limiting in nature. However, if this is taken as meaning that the intervention approach applied in that case was associated with a protective effect *ρ* < 1, this could have negative implications if this is used as a pretext for applying the same approach during an outbreak in which *R*_*s*_ >> 1. For example, data from the Diamond Princess outbreak have been used to inform interventions in other congregate settings.

Similarly, just as the generalizability of inferences about infection control are limited by the highly-selected nature of such analyses, these results also show the potential peril associated with extrapolating backwards from single outbreaks to characterize the transmission characteristics at a population level. Although it is possible that the average infectiousness among pre-intervention cases on the *Diamond Princess* cruise ship was nearly 15, as suggesed by Rocklöv et al. (4), our results show how even moderate amounts of heterogeneity can result in large overestimates of pre-intervention infectiousness, and can be characterized in large drops in the value of *R*_*s*_(*t*), even when the value of *R*_*s*_ is near the estimated population average value of 2-3 for SARS-CoV-2.

It important that these findings not be interpreted as an argument against the use of before-and-after comparisons as a tool for characterizing intervention effects. Often, in rapidly emerging and changing situations, they are the only tool at hand. Instead, we argue for a more statistically rigorous approach to accounting for the selection bias and heterogeneity which may give rise to these overly-optimistic inferences. For example, conditioning on the probability of observing an outbreak of size > *c* in the likelihood when fitting model parameters *R*_*s*_ and *k*_*s*_ and *ρ*_*s*_ would allow estimates to be adjusted for the observation bias at the center of these faulty inferences. One difficulty associated with this, however, and a weakness of the current analysis, is the the exact threshhold is typically unknown and is instead a function of the perceived severity of the outbreak among other factors that may be difficult or impossible to observe directly. Future analyses should examine whether the threshhold can plausibly be treated as a nuisance parameter that is either estimated or integrated out during the process of model fitting.

The COVID-19 pandemic has shown clearly that transmission models are critical tools for addressing and interrupting infectious disease transmission in real time. At the same time, the pandemic has also revealed the serious public health consequences of model misspecification and the critical need to have a suite of validated tools at the ready to guide intervention during future events. Future work should focus on directly addressing the out-of-sample predictive utility of such models, which is the acid test of their utility as tools for guiding real-world infectious disease surveillance and itnervention.

## Data Availability

Code to re-generate output will be made available upon request.

## Notes

### Competing Interest Statement

The authors have declared no competing interest.

### Funding Statement

JZ & NM were funded by an award from the Centers for Disease Control and Prevention

### Author Declarations

This study was conducted using only deidentified secondary data and not subject to IRB approval.

